# ICUBAM: ICU Bed Availability Monitoring and analysis in the *Grand Est région* of France during the COVID-19 epidemic

**DOI:** 10.1101/2020.05.18.20091264

**Authors:** Consortium ICUBAM, Laurent Bonnasse-Gahot, Maxime Dénès, Gabriel Dulac-Arnold, Sertan Girgin, François Husson, Valentin Iovene, Julie Josse, Antoine Kimmoun, François Landes, Jean-Pierre Nadal, Romain Primet, Frederico Quintao, Pierre Guillaume Raverdy, Vincent Rouvreau, Olivier Teboul, Roman Yurchak

## Abstract

**Background:** Reliable information is an essential component for responding to the COVID-19 epidemic, especially regarding the availability of critical care beds (CCBs). We propose three contributions: a) `ICUBAM` (ICU Bed Availability Monitor), a tool which both collects and visualizes information on CCB availability entered directly by intensivists. b) An analysis of CCB availability and ICU admissions and outcomes using collected by `ICUBAM` during a 6-week period in the hard-hit *Grand Est région* of France, and c) Explanatory and predictive models adapted to CCB availability prediction, and fitted to availability information collected by `ICUBAM`.

**Methods:** We interact directly with intensivists twice a day, by sending a SMS with a web link to the `ICUBAM` form where they enter 8 numbers: number of free and occupied CCBs (ventilator-equipped) for both COVID-19 positive and COVID-19- negative patients, the number of COVID-19 related ICU deaths and discharges, the number of ICU refusals, and the number of patients transferred to another *région* due to bed shortages. The collected data are described using univariate and multivariate methods such as correspondence analysis and then modeled at different scales: a medium and long term prediction using SEIR models, and a short term statistical model to predict the number of CCBs.

**Results:** `ICUBAM` was brought online March 25, and is currently being used in the *Grand Est région* by 109 intensivists representing 40 ICUs (95% of ICUs). `ICUBAM` allows for the calculation of CCB availability, admission and discharge statistics. Our analysis of data describes the evolution and extent of the COVID-19 health crisis in the *Grand Est* region: on April 6th, at maximum bed capacity, 1056 ventilator-equipped CCBs were present, representing 211% of the nominal regional capacity of 501 beds. From March 19th to March 31st, average daily COVID-19 ICU inflow was 68 patients/day, and 314 critical care patients were transferred out of the *Grand Est* region. With French lockdown starting on March 17th, a decrease of the daily inflow was found starting on April 1st: 23 patients/day during the first fortnight of April, and 7 patients/day during the last fortnight. However, treatment time for COVID-19 occupied CCBs is long: 15 days after the peak on March 31st, only 20% of ICU beds have been freed (50% after 1 month). Region-wide COVID-19 related in-ICU mortality is evaluated at 31%. Models trained from `ICUBAM` data are able to describe and predict the evolution of bed usage for the *Grand Est* region.

**Conclusion:** We observe strong uptake of the `ICUBAM` tool, amongst both physicians and local healthcare stakeholders (health agencies, first responders etc.). We are able to leverage data collected with `ICUBAM` to better understand the evolution of the COVID-19 epidemic in the *Grand Est* region. We also present how data ingested by `ICUBAM` can be used to anticipate CCB shortages and predict future admissions. Most importantly, we demonstrate the importance of having a cross-functional team involving physicians, statisticians and computer scientists working both with first-line medical responders and local health agencies. This allowed us to quickly implement effective tools to assist in critical decision-making processes.

## I. Introduction

In 5 to 10% of patients admitted for COVID-19, there are complications characterized by acute respiratory failure requiring admission to intensive care units (ICUs) with mechanical ventilation [Grasselli et al., 2020]. France has been been the 3rd most impacted country in Europe and the pandemic spread most quickly in the French *Grand Est région* (population of 5.5 million), beginning in the end of February. ICUs were quickly overwhelmed and reliable information on the availability and location of ventilator-equipped critical care beds (CCBs) quickly became essential for efficient patient and resource dispatching. Predicting the future load of the various ICUs also became necessary to better anticipate transfers and bed openings.

To meet these needs, a consortium including intensivists, computer scientists, statisticians and physicians developed `ICUBAM` ^1^ (Intensive Care Unit Bed Availability Monitor), which allows a network of intensivists to provide information in real-time on the capacity of their unit. This information can then be visualized on a map for use by intensivists and health authorities^2^. Supported by Inria (French national research institute for the digital sciences), `ICUBAM` is an open-source web-based application, available at GitHub and can be easily deployed elsewhere. `ICUBAM` is currently used by 130 ICU wards in 40 *départements*^3^, and inventories more than 2,000 ICU beds capable of accommodating a COVID-19 patient.

`ICUBAM`’s effectiveness results from the direct involvement of intensivists in the field who enter their ICU’s data directly into the system (see Appendix E for comparison of `ICUBAM` data with other administrative sources). Analysis of the collected data is used to monitor the burden on ICUs during the pandemic, and allows for anticipating CCB needs as well as for modeling the epidemic’s evolution.

This paper has three main contributions: In Section II, we present the open-source `ICUBAM` tool and demonstrate how it can be used by intensivists at patient’s bedside as well as by health authorities, to obtain reliable information on the availability of CCBs in the *Grand Est région* March 18t to April 30th. We provide a brief description of the tool and the data collected to highlight its ease-of-use and pertinence during the crisis. In section III, we present descriptive statistics and visualizations which testify to the importance of having real-time data during such a crisis. Finally, in section IV, we propose a SEIR model to describe the course of the pandemic using `ICUBAM` data, and show that it has better descriptive and predictive performances for patient inflow (number of patients admitted to ICU) and outflow (number of deceased and discharged ICU patients) than models calibrated on public data only or even on both sources of data. We complement this medium-horizon modeling by a short-horizon analysis using simpler statistical models to predict daily bed requirements.

## II. ICUBAM: a tool for bed availability monitoring

The `ICUBAM` application was built in response to the urgent need for intensivists to know real-time ICU bed availability over the *Grand Est* region. The first iteration was built in 3 days, and the system currently works as follows:

- Intensivists receive a text message 2 times per day (morning and evening)^4^ requesting bed information and containing a link to a web-based form.
- Physicians enter data into the form (fig. 1) and can also access the map (fig. 2, fig. 3) with the number of beds currently available for each *région* and ICU, as well as the ICUs’ contact information.

**Figure 1:**
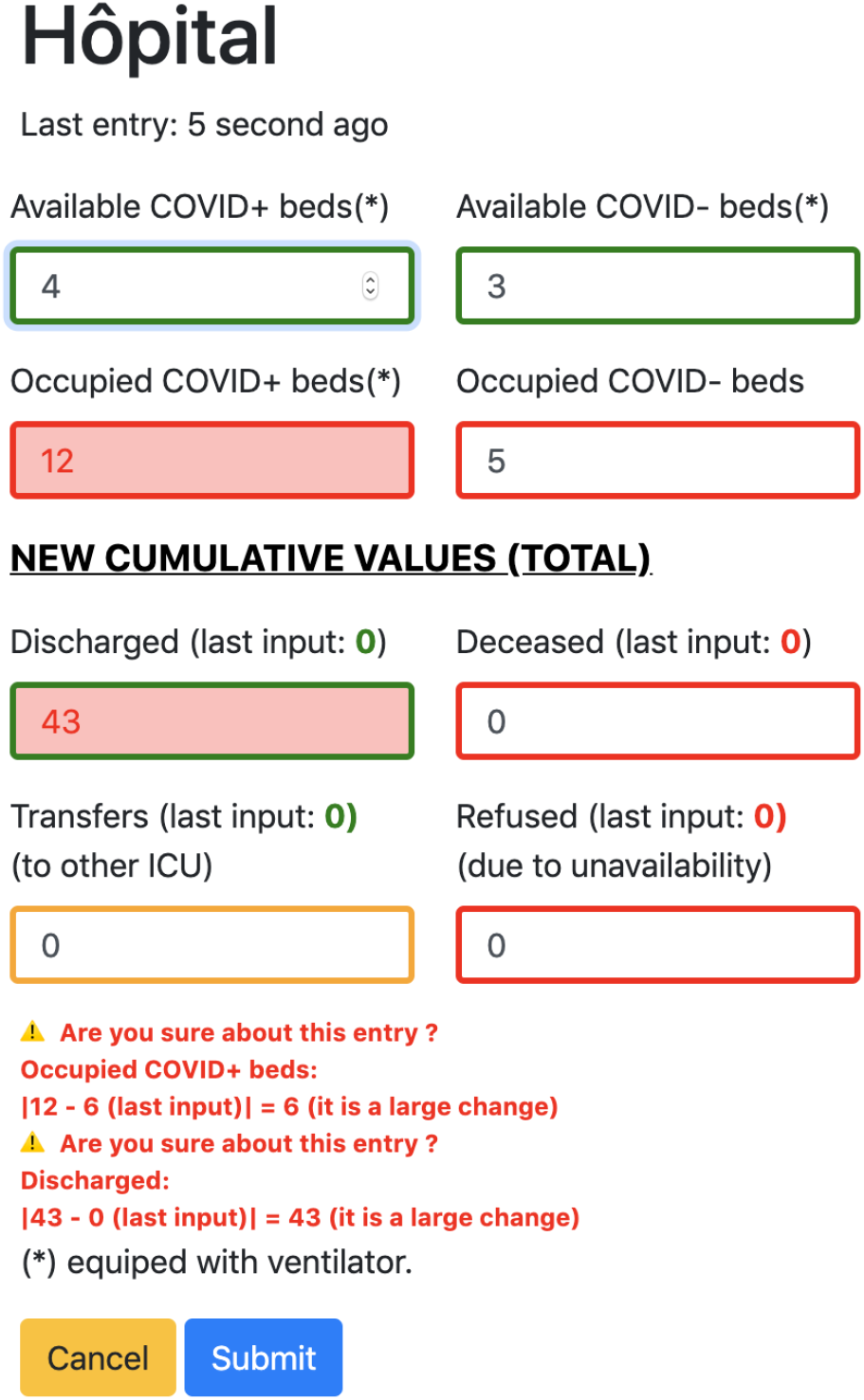
The form used by physicians to enter data into ICUBAM. Large or inconsistent entries trigger warnings. Previous values are pre-filled to encourage consistency.

**Figure 2:**
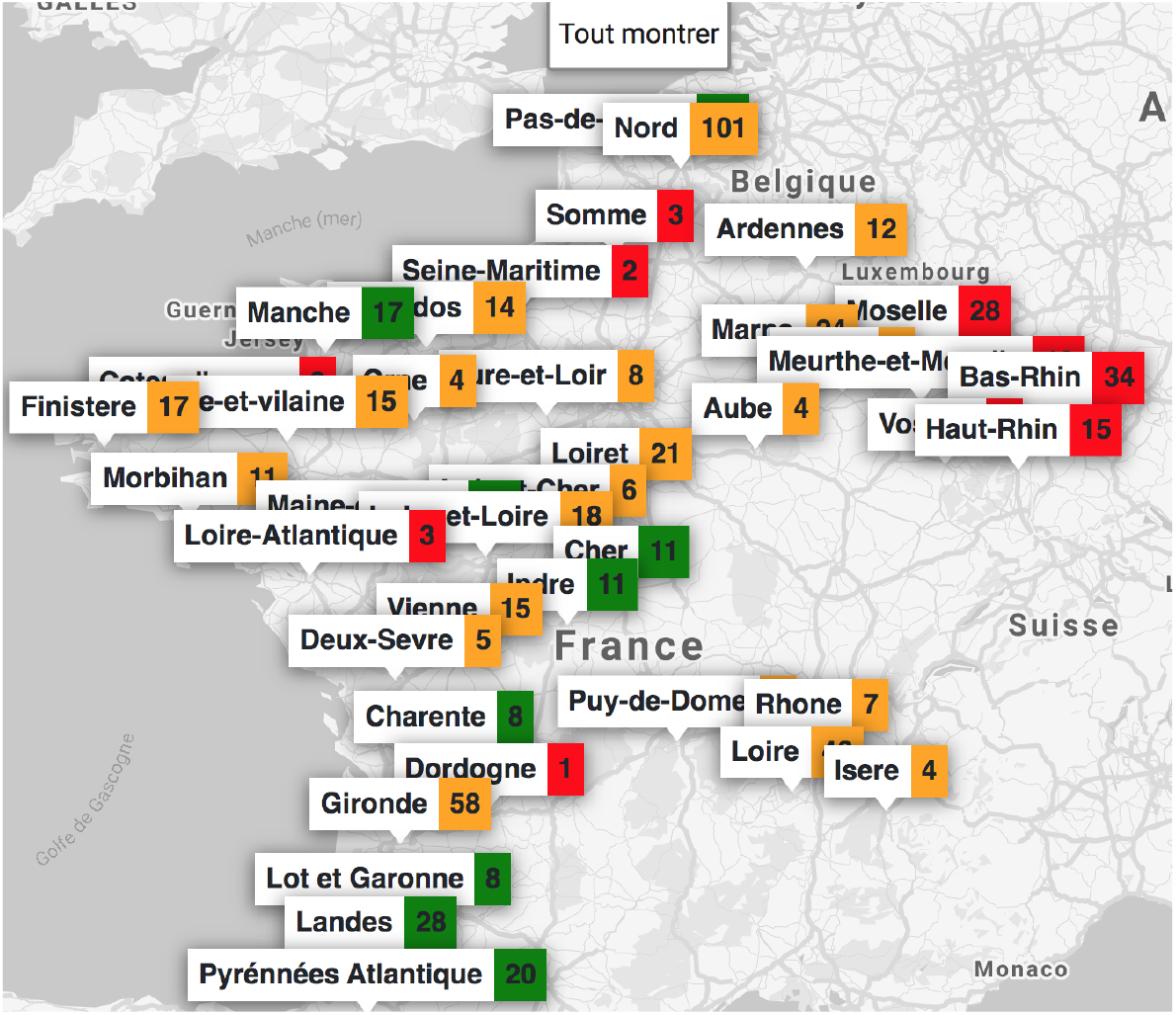
Map of available beds in France - April 18. Red indicates that more than 80% of the beds are occupied, orange between 50 and 80% and green less than 50%

**Figure 3:**
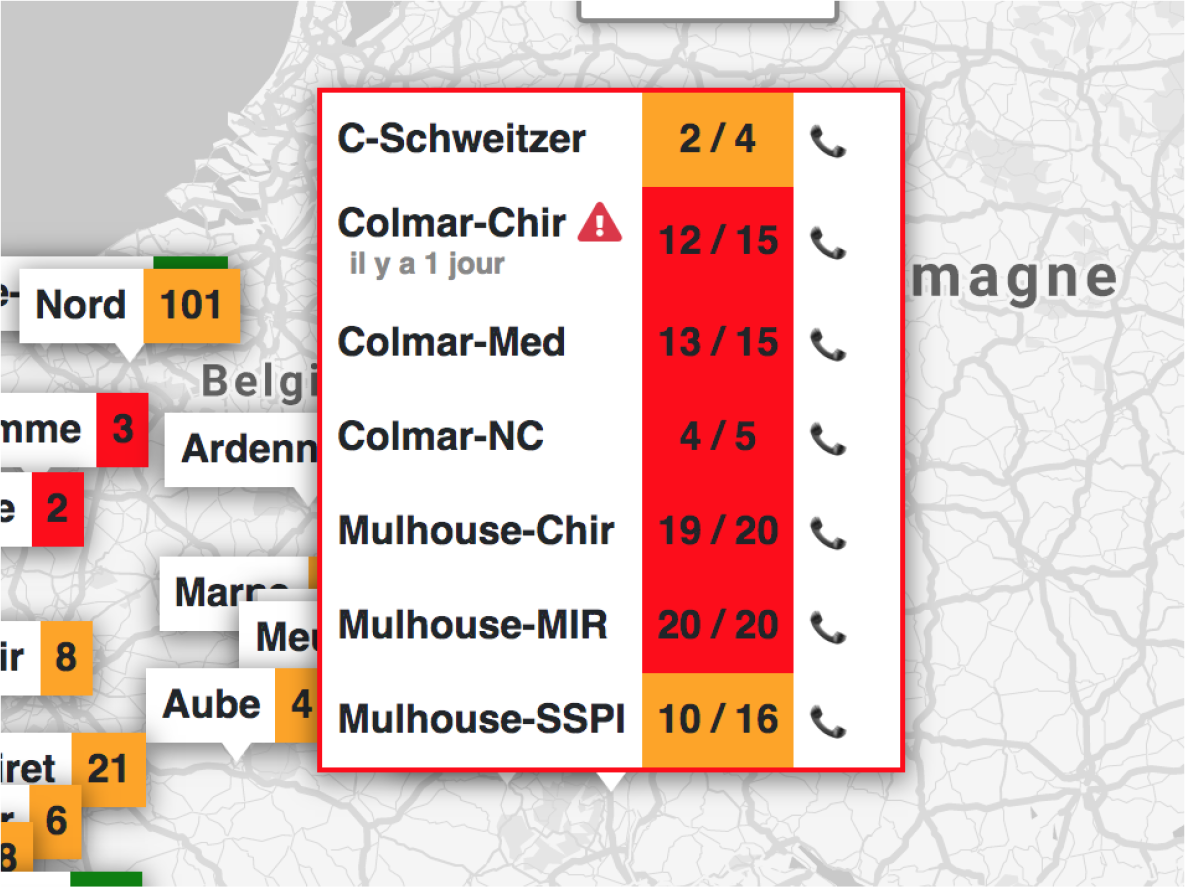
Number of beds occupied versus total number of beds in a *département*’s ICUs. The numbers reflect the last values indicated by the ICU. If the last input was more than a day ago, a warning is displayed to make it clear that the reported number might not be up to date.

Data entry can be performed in less than 15 seconds. The choice of variables in (fig. 1) was performed in collaboration with physicians from the *Grand Est* region, thus ensuring the relevance of the chosen statistics for both real-time use and downstream analysis. The 8 variables collected by `ICUBAM` allow study of the course of the pandemic on the ICU and are as follows: the number of free and occupied “COVID+” beds (in a COVID-19 floor), the number of free and occupied “COVID-” beds (in a non-COVID floor), the cumulative number of ICU-deceased COVID-19 patients, the cumulative number of ICU-discharged COVID-19 patients, the cumulative number of patients not accepted for entry due to lack of available CCBs and the cumulative number of patients transferred out of the ICU for capacity reasons (most often to other regions by medical trains). Some patients were also transferred to ICUs specialized in the management of refractory ARDS by VV-ECMO, which is identifiable when a transfer occurs while there are no refusals.

The national lockdown began on March 17th, and `ICUBAM` began operation on March 25. Partial data was manually collected beginning on March 18th and has been combined with the `ICUBAM` data. The analysis presented in this paper uses data until April 29th, and is performed at at the scale of a *département*.

`ICUBAM` was quickly adopted by 95% (40 out of 42) of hospitals in the *Grand Est région* and provided much-needed visibility to professionals on bed availability in their area. Authenticated accesses to map information are provided to regional health authorities. More details concerning the front and back-end of `ICUBAM` are provide in Appendix C.

## III. Visualization of data collected in the *région Grand Est* from 18th March to 29th of April

This section presents an analysis both of the patient flows & outcomes, as well as an analysis of ICU bed availability over the period of March 18th to April 29th.

### III.1. Patient admissions and outcomes

Data collected through `ICUBAM` during the study period allows us to understand patient admission and outcome dynamics for each ICU.

During the initial onset, it is crucial for health professionals and authorities to understand the pandemic’s evolution in order to react appropriately. This can be by public policy measures, but also by allocating resources and de-congesting saturated hospitals.

We can observe the evolution of the epidemic in the *Grand Est région* by following the evolution in admissions. Daily COVID-19 ICU admissions are presented in Figure 4. The number of patients entering is defined by

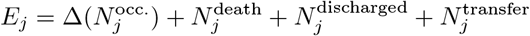

where *N_j_* is the quantity *N* on day *j*, and Δ(*N_j_*) represents the change in quantity *N* between day *j* and day (*j* − 1). The number of patients refused due to CCB shortages is not taken into account as they are often rerouted to an in-*région* ICU. Since April 1st, a decrease in the daily number of patients entering the ICU of the *Grand Est* is visible for all *départements*. This is most likely due to the lockdown measures put in place on March 17th, which suggests a two-week delay (mean duration between time from contamination to hospital admission) in system response to confinement measures [Zhu et al., 2020].

**Figure 4:**
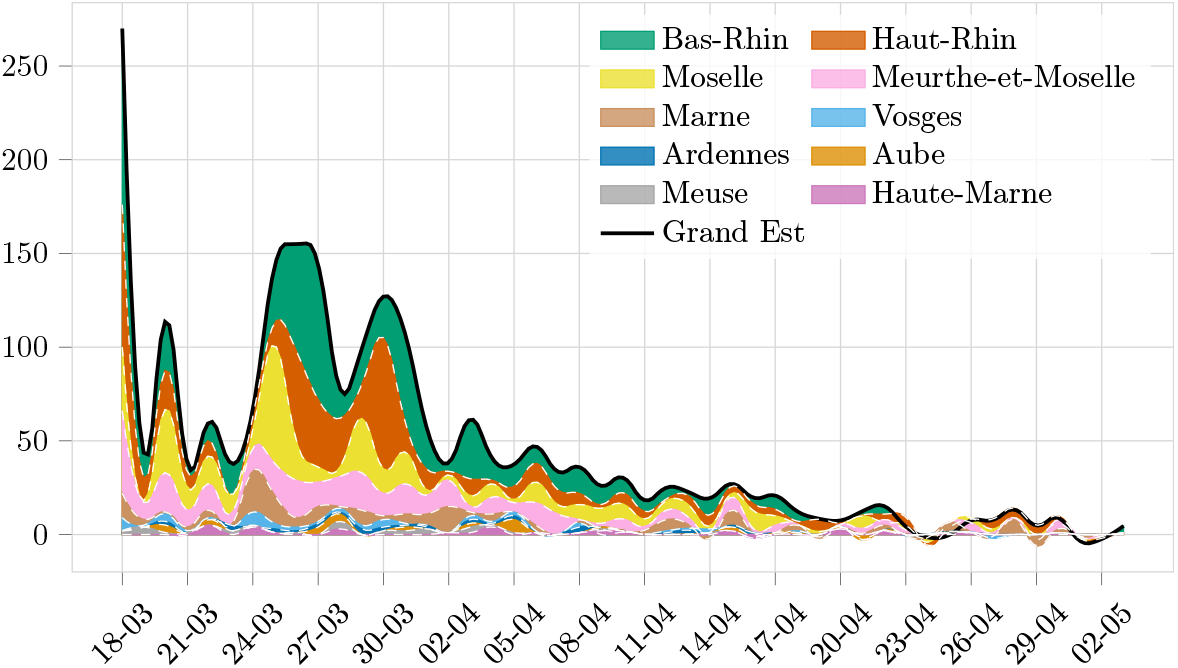
Daily number of COVID-19 ICU admissions for each *département* over the study period. Remark: the first value is disproportionate because it accounts for all patients of the period before the start of our data (March 18).

Patient outcomes are also important for understanding the situation. Cumulative deaths, discharges (living) and transfers from ICU are presented in Figure 5. In this Figure, we can see that both deaths and discharge statistics follow the same slope until March 27th, at which point the discharge rate increases while the death rate decreases. This might be due to an evolution in admission criteria of COVID-19 patients, as well as time-to-discharge being significantly longer than time-to-death. Details by *département* are given in Figure 14 in Appendix D.

**Figure 5:**
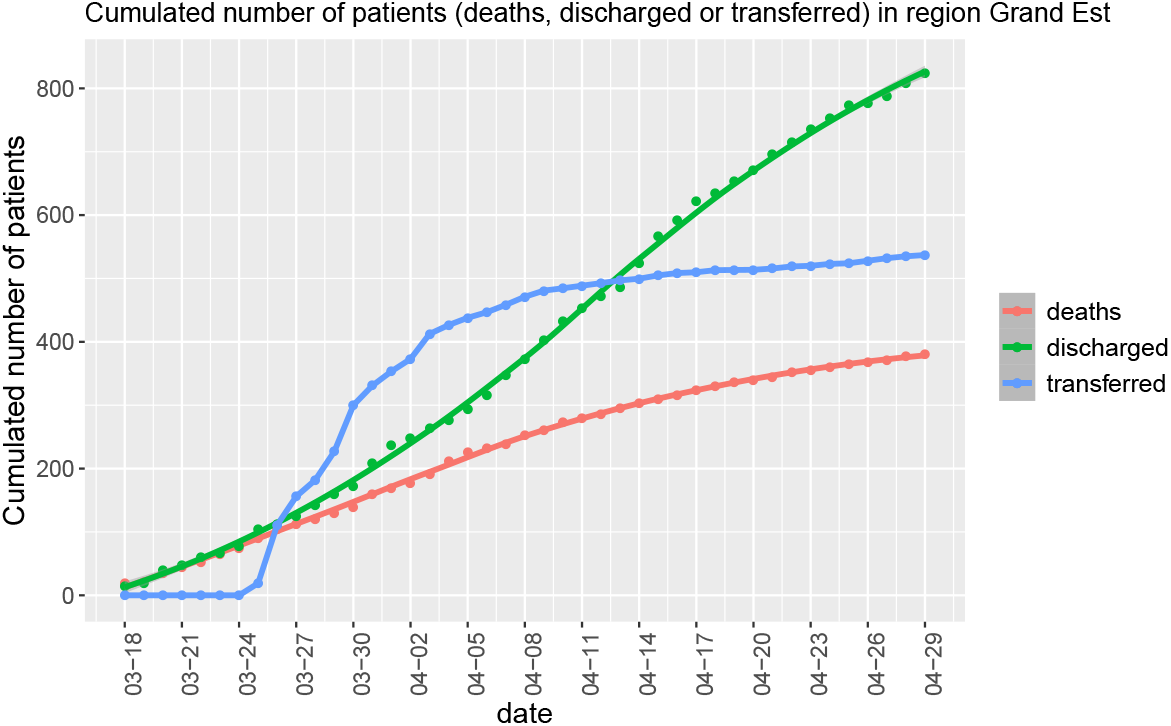
Cumulative value for patient deaths, discharges and transfers in the *Grand Est région* between March 18th and 29th of April. We can see that death and discharge rates are initially similar, but eventually discharges rate increases while death rate decreases.

Figure 6 visualizes patient outcomes during the study period for each *département*. In-ICU mortality 2 months after the onset of the pandemic’s start is 31.4%. This in-ICU mortality significantly lower than mortality reported by [Richardson et al., 2020]. Although the outcome of transferred patients is currently unknown, patients selected for transfer were generally more stable. One possible reason for this discrepancy is that ICU stay correlates strongly with outcome, and so patients likely to decease do so early on, whereas successful discharges happen much later. The longer time window of our study may therefore better capture the ultimate mortality rate of ICU stays.

**Figure 6:**
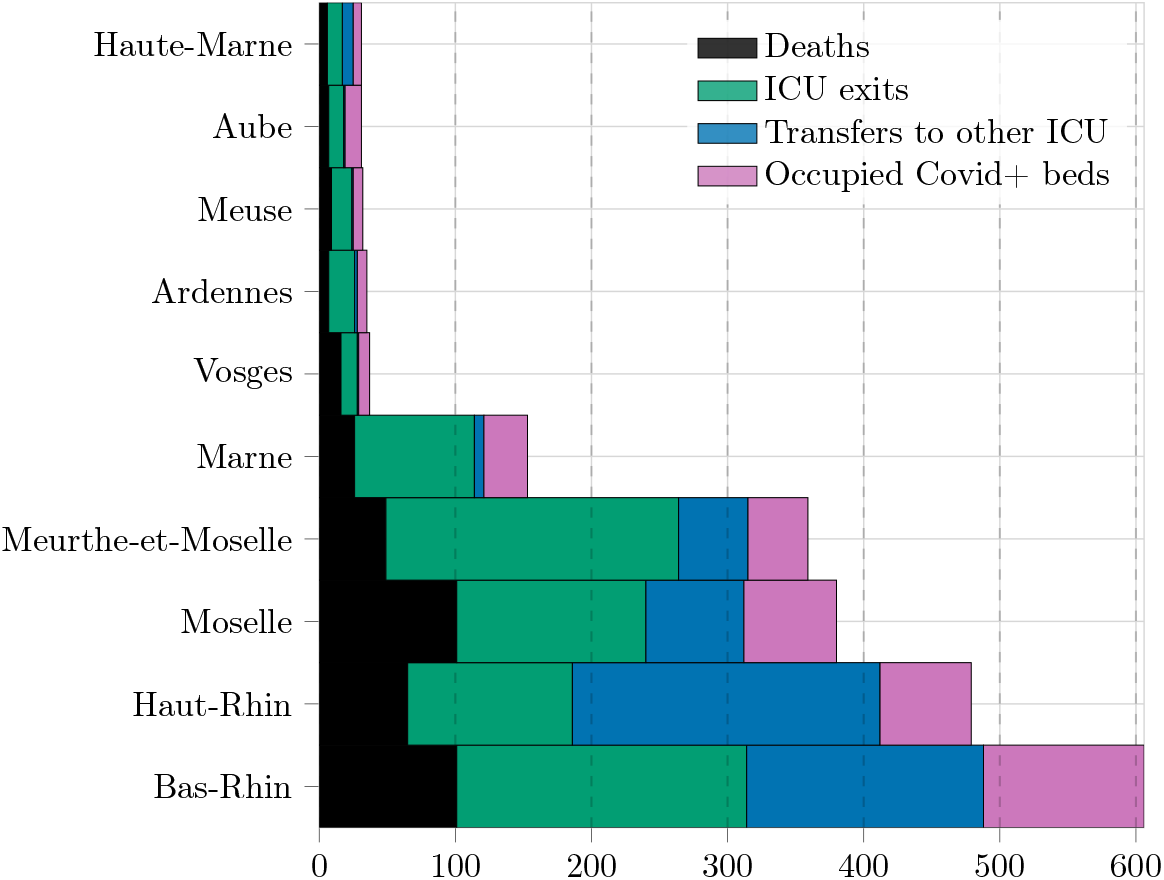
Summary of patient outcomes during the studied period for each *département*: total number of COVID-19-related deceased, discharged, or transferred patients as well as number of patients still in intensive at the end of the period. The sum of these four quantities represents the cumulative flow, over the period under consideration, of patients requiring admission to ICUs in each *département*.

### III.2. Evolution of supply and demand of CCBs

Figure 7 shows the evolving demand and supply of CCBs in the *Grand Est* region. This figure also demonstrates the impressive increase in CCB capacity, going from a nominal capacity of 501 beds to 1056 beds in 12 days i.e. 211% of nominal capacity. Nevertheless, this sudden increase in capacity was not enough to satisfy the need for CCBs, and almost 600 patients had to be transferred out of *région* to avoid surpassing the available CCB capacitys. After the pandemic’s apex, the decrease in the number of occupied CCBs is observable, but evolves slowly. COVID-19 CCB bed use went from 250 to 500 in 6 days (from 03/18 to 03/24), however the decrease from 750 to 500 has taken a total of 24 days (from 03/30 to 04/23), 4 times slower than the rate of admissions. This slow rate of discharges means that the system is still over-saturated, and a second wave of admissions would be difficult to absorb.

**Figure 7:**
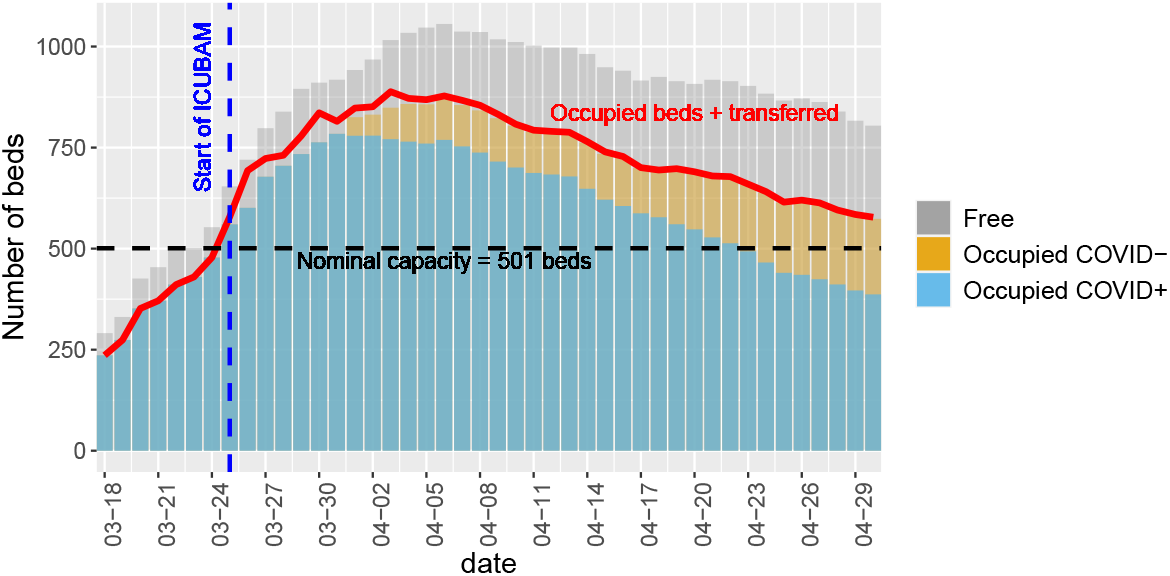
Number of beds occupied by COVID-19+ patients, non- COVID-19 patients, and total number of free ICU beds (regardless of COVID-19 status). The red curve represents the number of occupied beds plus the number of patients transferred to another region. Note that data before March 25th does not contain the number of transferred patients.

We can see that these region-wide evolutions are present in all individual *départements* through Figure 8. We have placed a dot to indicates the first date on which the nominal capacity is exceeded. The *Bas-Rhin* stands out, with a number of occupied beds which increased sharply on March 27. The number of people transferred was also significant at the time and started to decrease from March 30 (not represented here).

**Figure 8:**
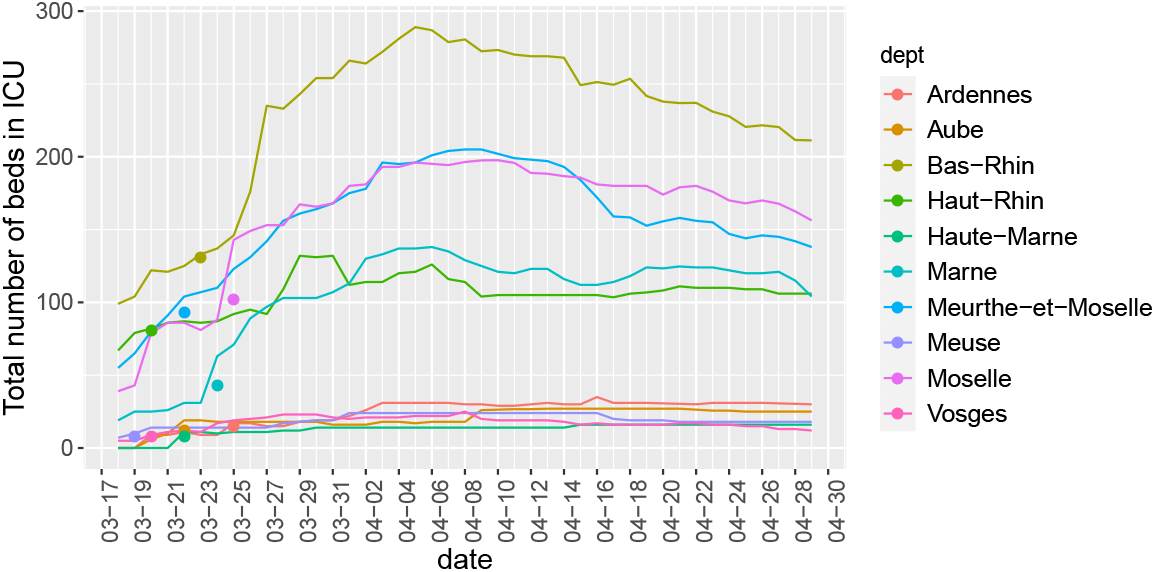
Evolution of the number of COVID+ CCBs for each *département*. The point indicates the first date on which the normal capacity is exceeded. Not all *département* have the same base population, but smaller *département* do not host many ICU beds.

As the epidemic evolved in the *Grand Est* region, different *départements* evolved differently in terms of CCB availability and demand. Temporal evolutions of each *départements*’ CCB occupation can be compared using a correspondence analysis [Husson et al., 2017, Lê et al., 2008]. Figure 9 can be interpreted as follows: two *départements* are close if they have the same evolution over time in terms of proportion of COVID+ occupied beds, and two dates are close if the distribution of occupied beds in the different *départements* is approximately the same. For the interpretation of the positions of a date and a *département*, we can say that a *département* goes towards the dates it is the most associated with, and away from dates it is the least associated with: a *département* is associated to a date if the number of occupied beds is high for this *département* at this date comparing to what happen in this *département* in the other dates, and also comparing to what happen at this date in the other *départements*. What is noticeable is that we can observe three clusters 3 of temporal behavior: between the 18 and 26 March (dates in red) the bed use increases significantly, between March 27th and April 14th (dates in orange) bed use is stable between *départements* and from April 15th to April 29th (dates in green) the number of beds start to go down.

**Figure 9:**
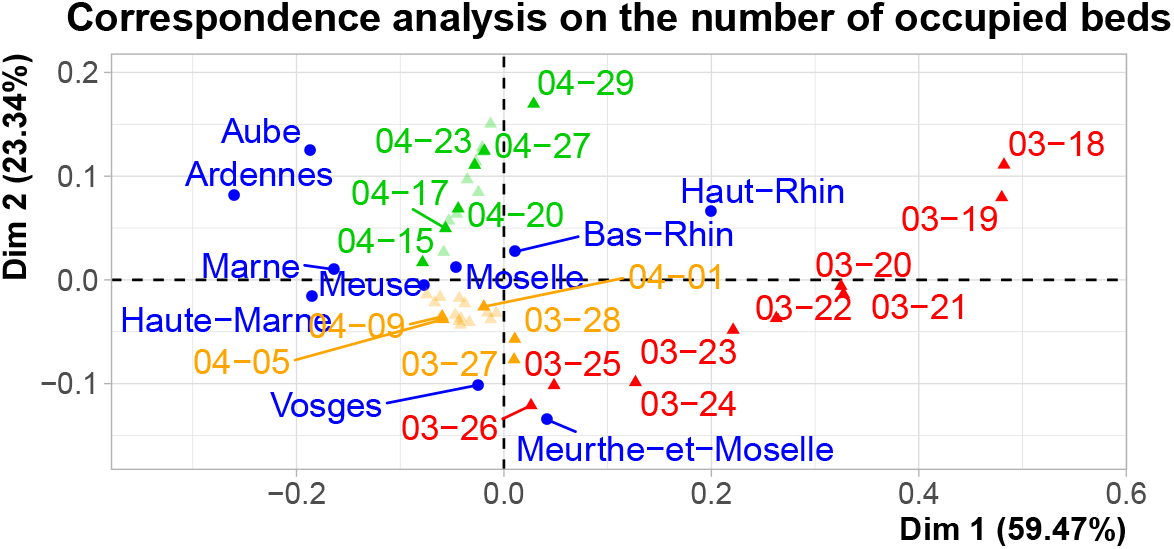
Correspondence analysis on the number of occupied beds per *département* and per day. This graphic presents both correspondences between dates, as well as *départements*, brought into the same plot. Some labels are not drawn.

The horizontal dimension naturally separates the *départements* in two: on the right side, *départements* such as *Haut-Rhin* (and to a lesser extent *Meurthe-et-Moselle*) that had many occupied CCBs at the beginning of the epidemic (i.e. dates in red from March 18th to 26th), and on the left side the other *départements (Ardennes, Aube, Haute-Marne, Marne*) that had relatively more occupied CCBs at the end of the period relative to the beginning. The vertical dimension creates a division between the *Meurthe-et-Moselle* and *Vosges départements* who reduce CCBs occupation earlier than other *départements*: they are at the opposite side of the green dates (beginning on April 15th) which suggests that their CCB occupation begins decreasing significantly earlier.

## IV. Modeling CCB availability

A key objective during the initial surge of patients is to properly allocate resources in a predictive rather than reactive manner, which requires predicting the number of critical care beds needed for each region. We therefore propose modeling the spread of the current pandemic using ordinary differential equations (ODEs) that are appropriate to describe the number of patients in ICUs and the number of cumulative deaths, as well as to provide a medium and long term prediction of CCB use. We also consider simple statistical models for short-term prediction of the number of released CCBs.

### IV. 1. Modeling the evolution of the pandemic with SIR and SEIR models

A large number of epidemiological models are available to describe the epidemic spread and the flows between the different states, either with the simplest SIR type model (susceptible, infected, recovered), or with more complex models, such as SEIR models (susceptible, exposed/incubation state, infected, recovered/deceased see e.g. [Hethcote, 2000, Allen, 2017]). These models can be made more complex to take into account the specifics of the particular epidemic and the available data. Many teams around the world are using official data to model the evolution of the pandemic at different scales in different countries. One can mention the resource center John Hopkins University [2020], the work of Lavielle [2020] which gives very good adjustments for the evolution of the pandemic at the scale of each country, and on French data, a study on the pre and post lockdown periods [Di Domenico et al., 2020].

For the *Grand Est* region, we have also found that it is possible to give a good account of the evolution of the epidemic based on public data, in each *département*, with classical models of the SIR or SEIR type. Another possibility is to combine public data with `ICUBAM` data (although the various sources are not always easy to align as illustrated in Appendix E). Nevertheless, we have obtained accurate descriptions and predictions of the number of patients in intensive care as described by `ICUBAM` data by modeling the underlying pandemic with a model calibrated on public data [Santé Publique France, 2020] for the number of hospitalized, discharged and deceased patients, and on `ICUBAM` data for CCB occupation. Indeed, the number of hospitalized patients is an additional element of data which is important to leverage. We may expect that the global dynamics of the number of hospitalized patients will constrain in the fits the one of the ICU patients. Somewhat surprisingly, we found that a model using only `ICUBAM` data gives as good results as more complex models based on both datasets. This also means that we get as good or better results with a smaller number of parameters, which suggests better predictive power for the purely `ICUBAM`-based model. For this reason, we present here the `ICUBAM`-only results.

Figure 10 illustrates the model used. It is a simple SEIR model which assumes that the different *départements* are independent (the data are collected after lockdown), and uses the number of patients admitted to ICUs and the number of exits (discharged and deceased patients), but considers a lengthy time-to-exit by adding a period before exiting *(ICU*_1_ *→ ICU*_2_ *→* exit) to account for the time of recovery (or death). More details are given in Appendix F. We calibrate the model parameters in order to have the best agreement between the observed data and the model outcome (with a maximum likelihood criterion). The proposed family of models have predictive capabilities as shown in Figure 11 and in Figure 17 in Appendix F. The modeling allows us to see a global trajectory and the coherence of the observations in relation to the evolution of the pandemic. For example, for the *Meuse département* in Figure 17 we can see that the observations ‘catch up’ with the model’s predictions.

**Figure 10:**
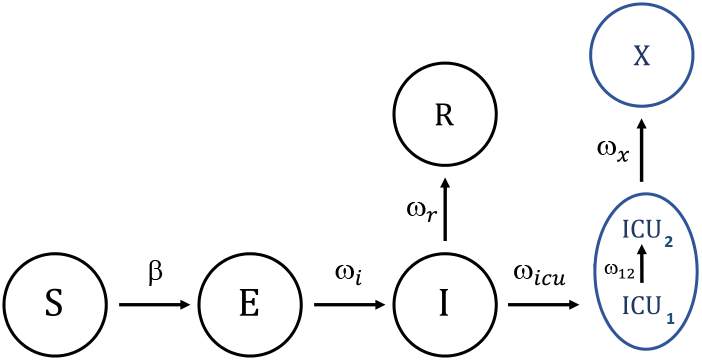
Flow shart of the SEIR type model with incubation compartment. In blue, the parts corresponding to the ICUBAM data on which the model is calibrated: number of patients in ICU, cumulative number of patients leaving the ICU (X for eXit, discharged and deceased patients).

**Figure 11:**
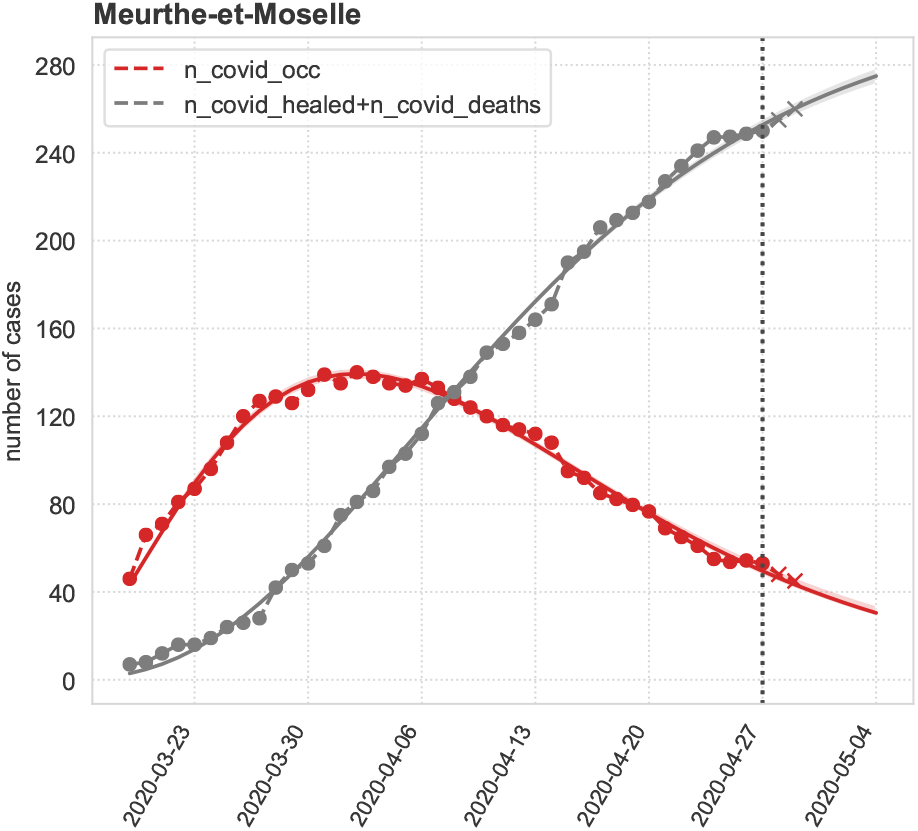
Fit a SEIR model for *département Meurthe-et-Moselle* using data up to the 27 of April (vertical dotted line; data: circles) and with prediction for the following days (crosses: data on 28 and 29 of April). Model ’susceptible/exposed/infected/ICU/exit’, with recovery (or death) period. Red: number of patients currently in ICU. Grey: cumulative number of patients either deceased or discharged from intensive care units.

However, predicting the number of released CCBs faces several problems. Models of the SIR type are well suited to account for the current number of patients in intensive care and the cumulative number of discharged and deceased patients. The number of released CCBs is calculated as the difference between the change in each of these numbers between two consecutive days. For large numbers, the differences will vary greatly for small variations in the estimated quantities. For a fit calibrated on data up to 26 April, for example, a number of CCBs released during the two following days, 27 and 28 of April, is predicted which is correct for the *départements Haut-Rhin* (9, observed 6), *Marne* (7 or 8, observed 10), *Moselle* (13 or 14, observed 11), but gives a large difference for the *Bas-Rhin* and *Meurthe-et-Moselle*. This can be understood by looking at figure 17: the predicted trajectory deviates slightly from the data, with the formation of quasi plateau that a SIR model cannot account for. A more refined modeling of the dynamics of the evolution of the patients’ condition - resulting in a wide distribution of ICU residence times, as discussed below - could be incorporated into the model.

### IV.2. Modeling the number of beds released with statistical models

To get short-term (per day) and refined prediction of the number of released CCBs (either due to death or discharge), we consider disconnecting the prediction of the number of admissions, which depends on the evolution of the pandemic, to the prediction of the number of beds released.

We evaluated several types of statistical models that leveraged number of ICU admissions in the preceding days as explanatory features. We ran regression models with variable selection, random forests, but also the simple average of the number of ICU admissions between 1 and 20 days before the date for which we want to predict the release. This latter simple method comes from the observation that the distribution of the number of days with invasive mechanical ventilation times is not significantly different from a Uniform distribution (see figure 12). Note that the data is obtained from a single hospital with a hundred of patients and should be refined. We also add, as a benchmark, the naive method that predicts the number of critical care beds released by the last observation of the number of critical care beds released.

**Figure 12:**
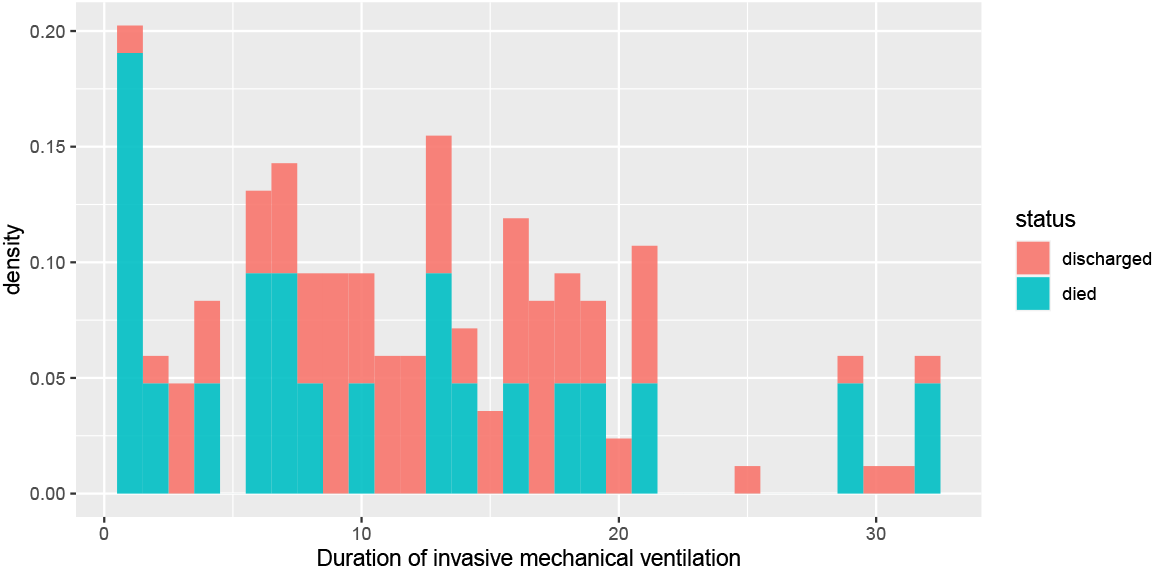
Duration of invasive mechanical ventilation before death or discharge.

To evaluate the models, we define a training set of all data until a date (called “Last training day” in Table 1) and then we calculate the predictions for the 2 days after. Table 1 gives the mean absolute error of prediction of the number of beds that would be released the next two days for each *département* when the models are trained using data until the last training day (1st column). It turns out that the average method gives the smallest errors (2.03) and improves significantly upon the last observation carried forward method (day before).

**Table 1:** Mean absolute error of prediction between observed values and predicted values given by four models: linear model (lm), random forests (RF), the average of inputs between D-1 and D-20 and the entry observed the day before; models learned with data from the 18 of March until the last training day (1st column) and predict for the 2 days after.

| Last training day | lm | RF | average | day before |
| --- | --- | --- | --- | --- |
| 2020-04-28 | 1.19 | 1.49 | 1.35 | 3.44 |
| 2020-04-27 | 1.81 | 1.77 | 2.19 | 3.71 |
| 2020-04-26 | 3.19 | 2.57 | 2.18 | 2.78 |
| 2020-04-25 | 3.65 | 3.22 | 2.61 | 2.30 |
| 2020-04-24 | 3.54 | 3.23 | 2.44 | 4.65 |
| 2020-04-23 | 3.72 | 2.27 | 1.80 | 2.85 |
| 2020-04-22 | 2.64 | 1.77 | 2.22 | 3.52 |
| 2020-04-21 | 2.04 | 1.17 | 1.49 | 2.75 |
| Mean | 2.72 | 2.19 | 2.03 | 3.25 |

If we wanted to predict at D+5, the method with the average remains the best since the mean of the error (last row of the table) would be 8.76 for linear model, 5.44 for random forest, 3.53 for the average and 9.30 for the last day.

We can also predict the number of exits for May 1 and 2 (see table 2).

**Table 2:** Prediction of the number of exits (sum of deaths and discharged) for May 1st and 2nd obtained with the linear model (lm), random forests (RF) and the average of the ICU admissions between D-1 and D-20; the models learned with the data until April 30.

|  | lm | RF | average |
| --- | --- | --- | --- |
| *Ardennes* | 0 | 1 | 1 |
| *Aube* | 0 | 1 | 0 |
| *Bas-Rhin* | 1 | 3 | 2 |
| *Haut-Rhin* | 4 | 5 | 5 |
| *Haute-Marne* | 1 | 1 | 1 |
| *Marne* | 2 | 4 | 3 |
| *Meurthe-et-Moselle* | 1 | 2 | 4 |
| *Meuse* | 0 | 1 | 0 |
| *Moselle* | 3 | 4 | 5 |
| *Vosges* | 0 | 1 | 0 |

## V. Conclusion

### V.1. Quality data, a flexible processes and an inter-disciplinary team

The great strength and particularity of `ICUBAM` is to be supported by an inter-disciplinary team of intensivists, engineers, researchers, statisticians, physicists and computer scientists who, together, designed and built the entire pipeline, from data collection to analysis, and communication of results in real-time to meet operational needs in an emergency context.

Data quality is a key challenge in the management of this crisis and the flexibility of the data collection as well as the direct contact with stakeholders to get and exploit important information (such as time spent in critical care) is essential.

Nevertheless, `ICUBAM` data, although granular, provide only a partial view of the pandemic as data were only collected for critical care beds. In addition, the strength of this tool is to collect data directly from the intensivists but it also implies that the data are necessarily limited: only the data that is immediately useful for intensivists are entered, and it is obviously not possible to increase their workload by asking them to enter more information - the lightweight interaction with our systems was key in having be used by so many ICUs.

The customized nature of `ICUBAM` also allowed it to be easily adapted as new needs arose (addition of a non-COVID ICU map, displaying age of the data, live plotting for physicians and health agencies).

### V.2. Impact of ICUBAM

From direct discussions with intensivists from the *Grand Est* region, `ICUBAM` has aided in disseminating information along two important axes: First, horizontally amongst other intensivists the *Grand Est* region. `ICUBAM` quickly gained traction amongst these front-line physicians by creating both visibility of the situation in nearby ICU wards, but also by creating an important information connection between private and public-sector ICUs to easily share their availability in a unified platform.

Secondly, `ICUBAM` has proven to be a useful tool for sharing information vertically from the physician-level, and up through authorities. This distribution of information from `ICUBAM` hopefully contributed to the balance between demand and availability of critical care beds in the *Grand Est* region.

From a public and clinical health perspective, better understanding of the epidemic’s mechanisms and better planning of resource needs and triage of critical care patients can have a substantial impact on patient care and possibly save lives. We hope that `ICUBAM` can be useful to assist the decision-making process by providing a framework to collect and analyze detailed and reliable data, and that the analyses provided herein give some insights on how COVID-19 can quickly overload even a well-structured health system.

## Data Availability

The data are available on request

https://icubam.github.io/

## Acknowledgements

Thanks to all intensivists from *Grand Est région* who made this study possible.

Thanks to the French Intensive Care Society for enthusiastically supporting `ICUBAM`, the ARS *Grand Est* and the health authorities who, despite the complex context, have allowed us to work on `ICUBAM`.

Thanks also to Professor Catherine Paugam Burtz who put the main actors of the project in contact as well as to Dr Sacha Rozencwajg.

Thanks to Achille Thin, Geneviève Robin, Romain Egele for their help at the start of this project, and Eric Mermet for his help in the making of maps. Thanks to Thomas Merckling PhD from INSERM CIC-P 1433 for his contribution to the prototype of `ICUBAM`.

We would also like to warmly thank Inria who supported us during this period and a special thanks to Hugues Berry and Alexandre Gramfort.

## Author contributions

The authorship of this article is in alphabetical order. The contributions are as follows:

- Project design and organisation: GDA, JJ, AK and OT
- Creation of the ICUBAM interface: GDA, AK, VI, OT
- System: GDA, SG, RP, FQ, PGM, VR, OT, RY
- Data processing: GDA, VI, AK, FL, RY
- Descriptive statistics: GDA, FH, JJ, AK, VI, FL
- Modeling: LBG, FH, JJ, AK, JPN
- Writing: GDA, FH, JJ, AK, JPN

All authors reviewed the manuscript.

The instance of ICUBAM used in this paper runs on Inria servers. Correspondence and requests for materials should be addressed to.

We declare on our honor that there are *no ethical approval* questions related to this manuscript; there are no involvement of humans. In accordance with the Common Rule, review by the institutional review board and informed consent was not required for this study due to the non-human participant research determination. Inria’s legal department guarantees that project is in accordance with the General Data Protection Regulation and that data are transferred to the Ministry of health.

The ethics board from french intensive care society gave ethical approval for the paper and project.

In accordance with the Common Inria’s legal department guarantees that project is in accordance with the General Data Protection Regulation and that data are transferred to the Ministry of health.

## Code availability

`ICUBAM` is open-source and is available at https://github.com/icubam/icubam/. All source code used for analysis and modeling will be made available on GitHub.

Code for statistical analysis and modelling is written in R.

Code for the SIR type modeling is written in Python. To compute the credible regions (see the Appendix) we made use of the PyMC3 package for Python [Salvatier et al., 2016] (https://docs.pymc.io/).

## Appendix

### A. Genesis of ICUBAM

The project is the result of a personal initiative by an Antoine Kimmoun M.D., and intensivist from the *Grand Est région* of France who identified the urgent need to visualize occupied COVID-19 CCBs in a real-time manner. He started to develop a prototype on March 18, 2020 to collect the data on the availability of beds by placing phone calls and centralizing the information in a spreadsheet. `ICUBAM` development began on Sunday March 22, 2020 after a meeting between Antoine and team of engineers and researchers from Polytechnique, Inria, and elsewhere. On Wednesday March 25, 2020, we launched `ICUBAM` in the *Grand Est région* in agreement with the ARS (Regional Health Agency). Other regions quickly started using `ICUBAM` (Center Val-de-Loire, Brittany, AURA, New Aquitaine, etc.). `ICUBAM` is currently used by 130 ICUs in 40 *départements*, which represents more than 2,000 COVID-19 CCBs.

### B. French administrative divisions - The *Grand Est région*

The three main French administrative divisions are: the *commune* (municipality); at a larger scale the *département* (96 *départements* in Metropolitan France, labelled from 1 to 95, with 2A and 2B for Corsica); and the *région* (12 in Metropolitan France, excluding Corsica), aggregating several neighbouring *départements*. The typical diameter of a *département* is 100 km, and the one of a région, 250 km. The Grand Est région, in the North-East part of France, is composed of the following *départements*: *Ardennes* (08), *Aube* (10), *Marne* (51), *Haute-Marne* (52), *Meurthe-et-Moselle* (54), *Meuse* (55), *Moselle* (57), *Bas-Rhin* (67), *Haut-Rhin* (68), *Vosges* (88).

### C. icubam open-source software

#### C.1. User flow

The user’s journey on `ICUBAM` has been thought with physicians to ease their way through finding quickly bed availability nearby, under the particular stress constraints witnessed in pandemic times.

It starts with a text message (usually SMS, but other means are also supported) sent to the user on a regular schedule. This messages contains as token, that is generated specifically for a given user of a given ICU. `ICUBAM` also supports automatic refreshing of tokens periodically.

This link containing the token is needed to access the the availability form in which the user can easily enter the 8 counts described in this document. To prevent typos, big changes in number from an update of those counts to the other triggers a warning, inviting the user to double check the values.

Having entered the current state of bed availability in their ICU, the users are eventually redirected to the map page, where color codes and warning signs helps the physician narrowing down his search of bed availability to nearby ICUs.

The user’s journey ends up with one or several call to some ICUs, directly from the map.

#### C.2. Architecture

`ICUBAM` is divided into four web services: communication with the database, the web interface for physicians, a back office web interface for administrators and a message scheduler service.

#### C.3. Dashboards

A dashboard (fig. 13) is available for some users with granted access through the `ICUBAM` backoffice, which is a web interface to manage `ICUBAM`’s ecosystem. It contains summary statistics (appendix C.4) and some of the plots presented in this very document.

**Figure 13:**
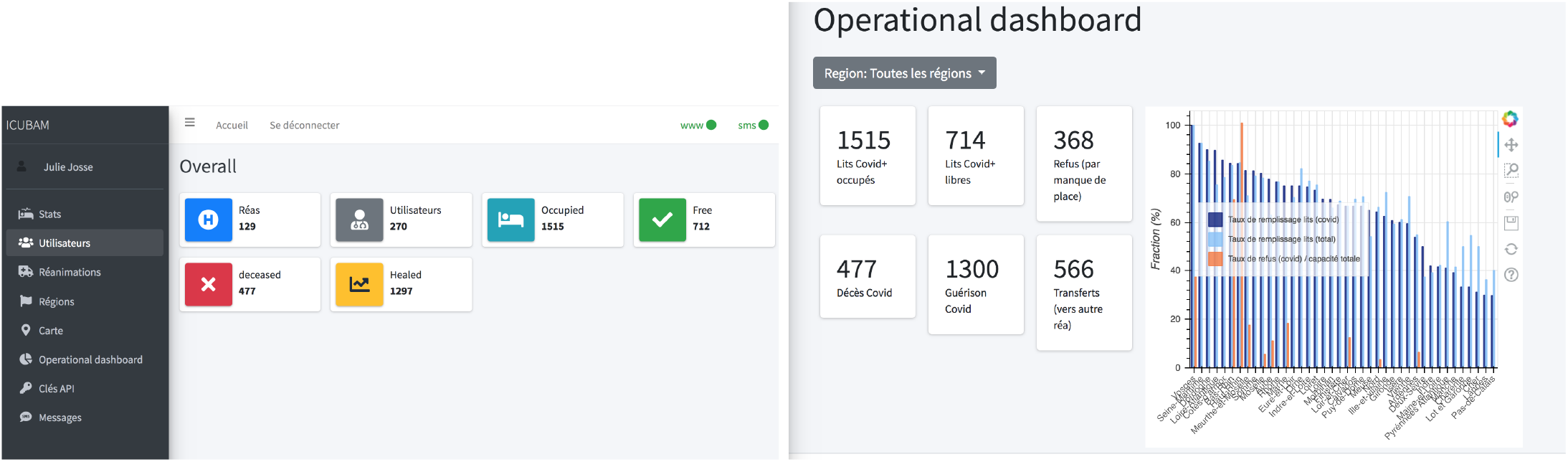
Dashboard for medical doctors: Excerpt from the April 18 dashboard.

**Figure 14:**
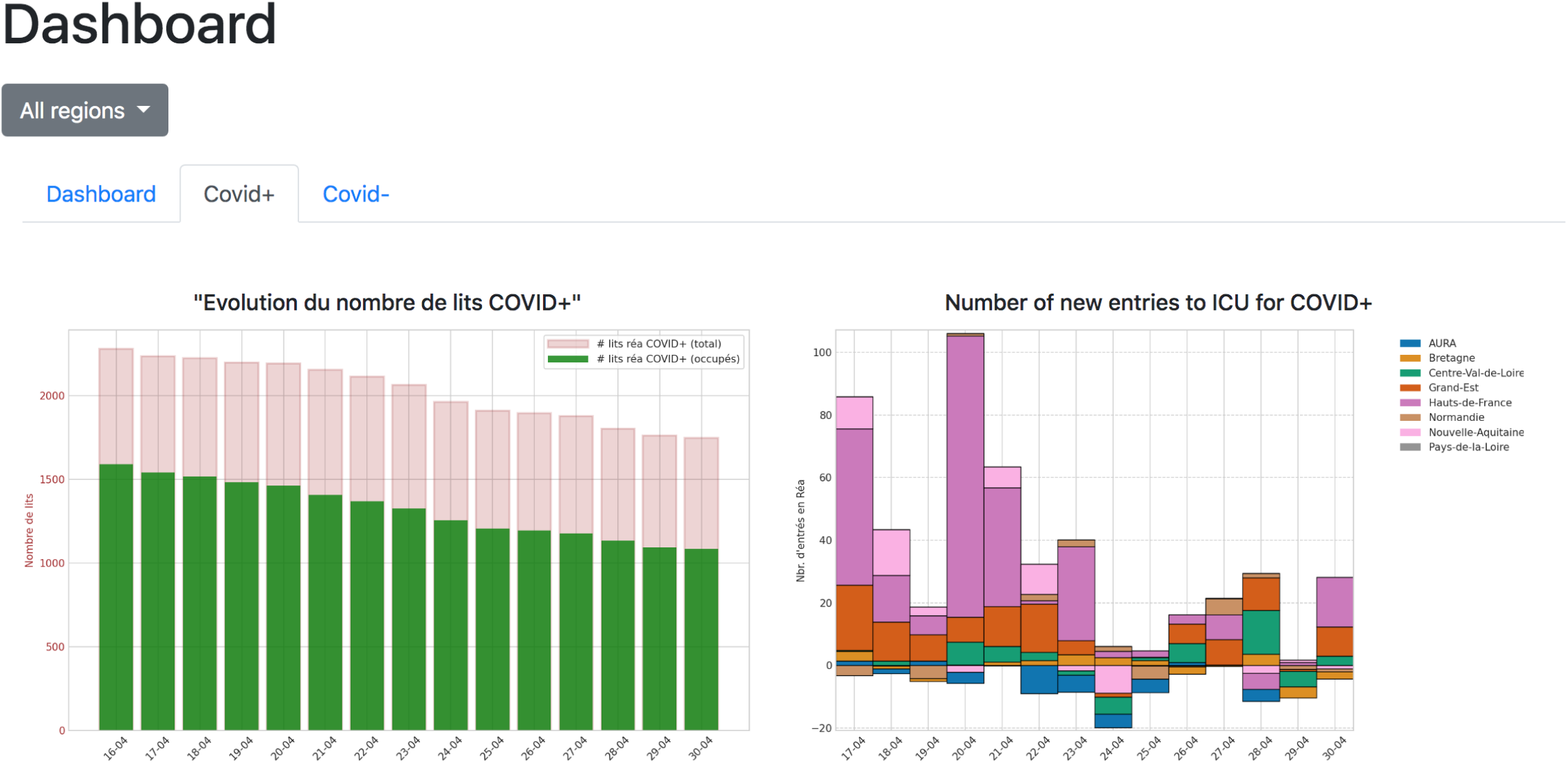
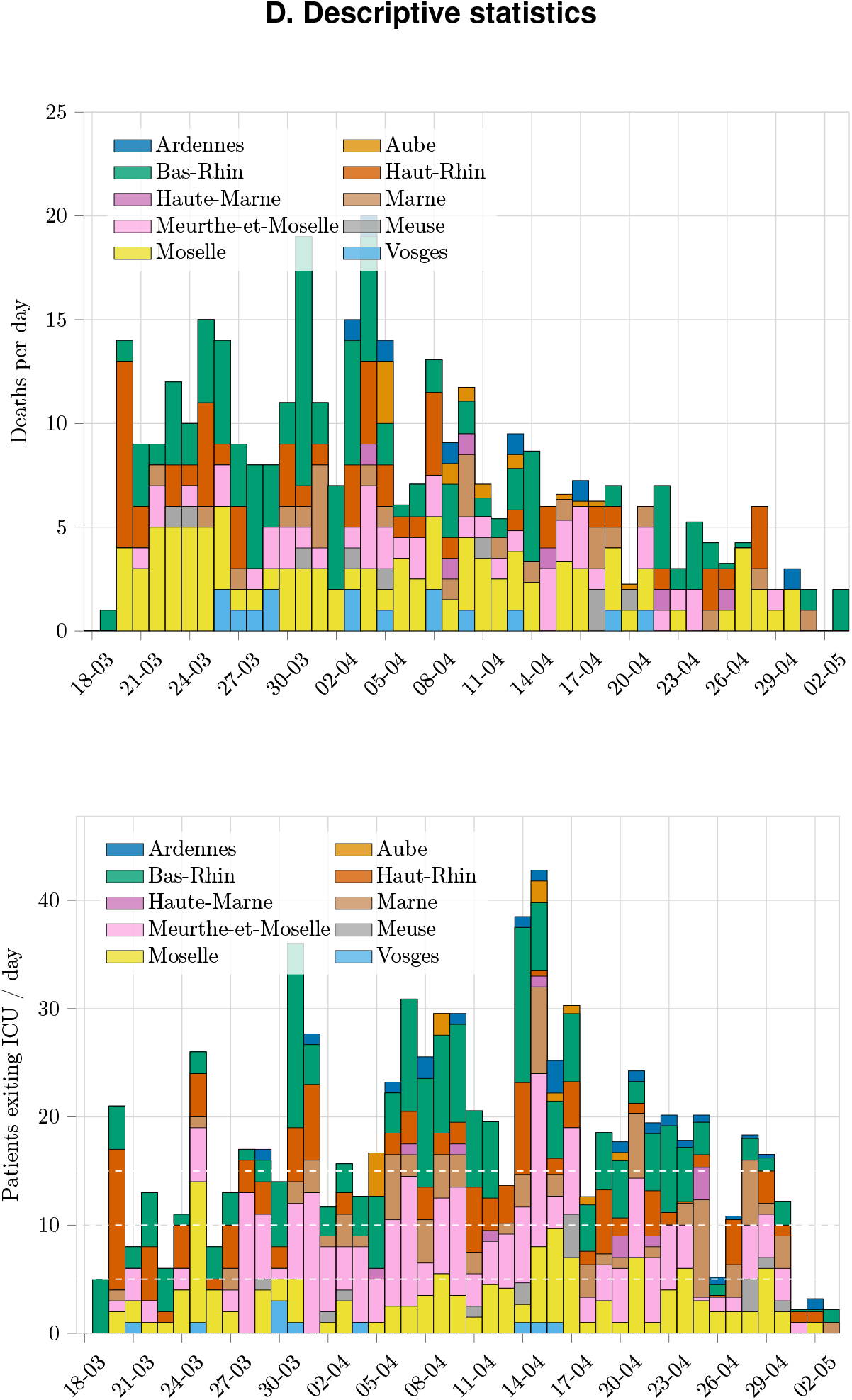
Daily number of deaths in ICU (top) and ICU’s discharged (bottom) per *département*.

#### C.4. Data

Note that the data collected for this study is done so by an instance of ICUBAM hosted by Inria and not part of the open-source software. External developers as well as external deployments of ICUBAM have no access whatsoever to this data, but do own their own data. This separation between data and software guarantees both data security and reproducibility of the ICUBAM service outside of the French context.

### D. Descriptive statistics

#### E. Comparison between ICUBAM and official public data

The analysis of `ICUBAM` data and the use of public data [Santé Publique France, 2020] has also highlighted the difficulty of comparing the data collected, which often represent different realities. For example, `ICUBAM`’s scope only covers resuscitation beds equipped with a ventilator, whereas very often critical care beds include equipped or not with ventilators. However, this implies that the number of ICU patients should be larger in the public data case, which is not always the case, as can be seen in the right panel of fig. 16.

**Figure 15:**
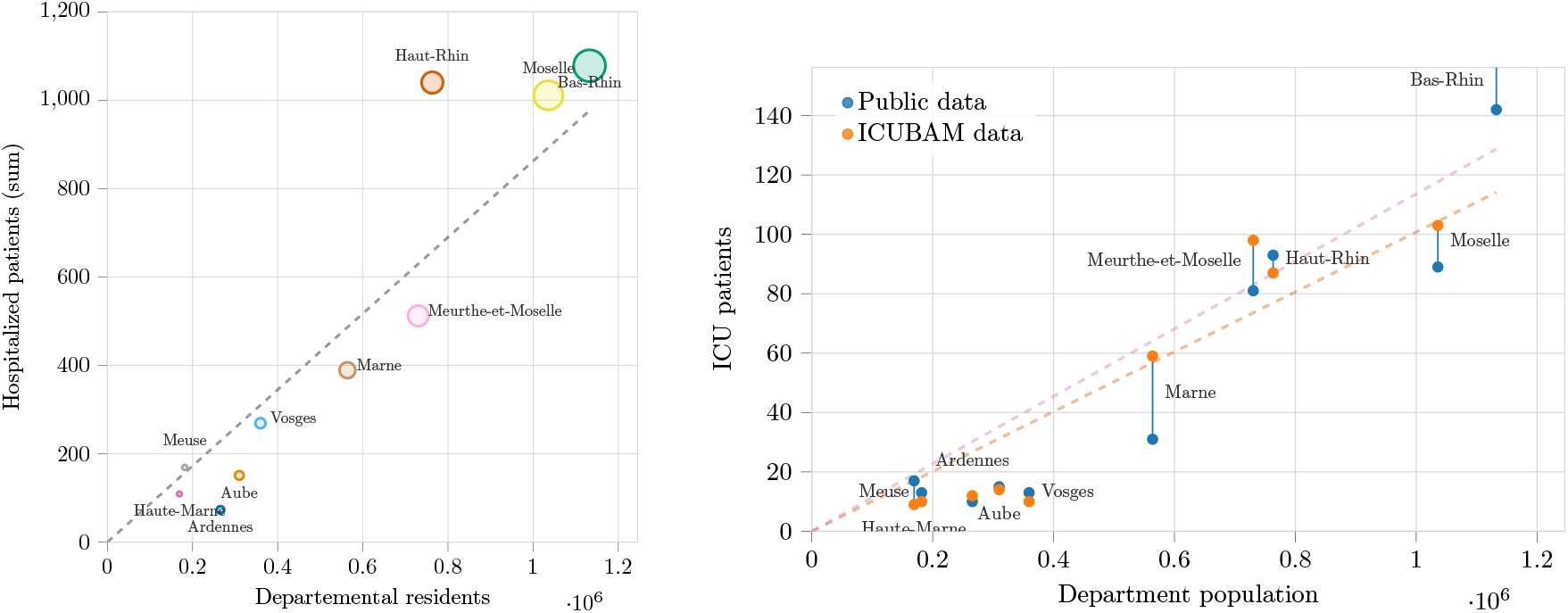
**Left**: total number of patients (COVID or not) hospitalized on 29th of Aprilin each *département*, based on official public data (source Santé Publique France [2020]). The slope of the regression line was calculated by excluding the *Haut-Rhin* (outlier) and the intercept was set at 0 (a null population implying a null number of hospitalized patients). **Right**: comparison between official public data and `ICUBAM` data on the number of ICU cases.

**Figure 16:**
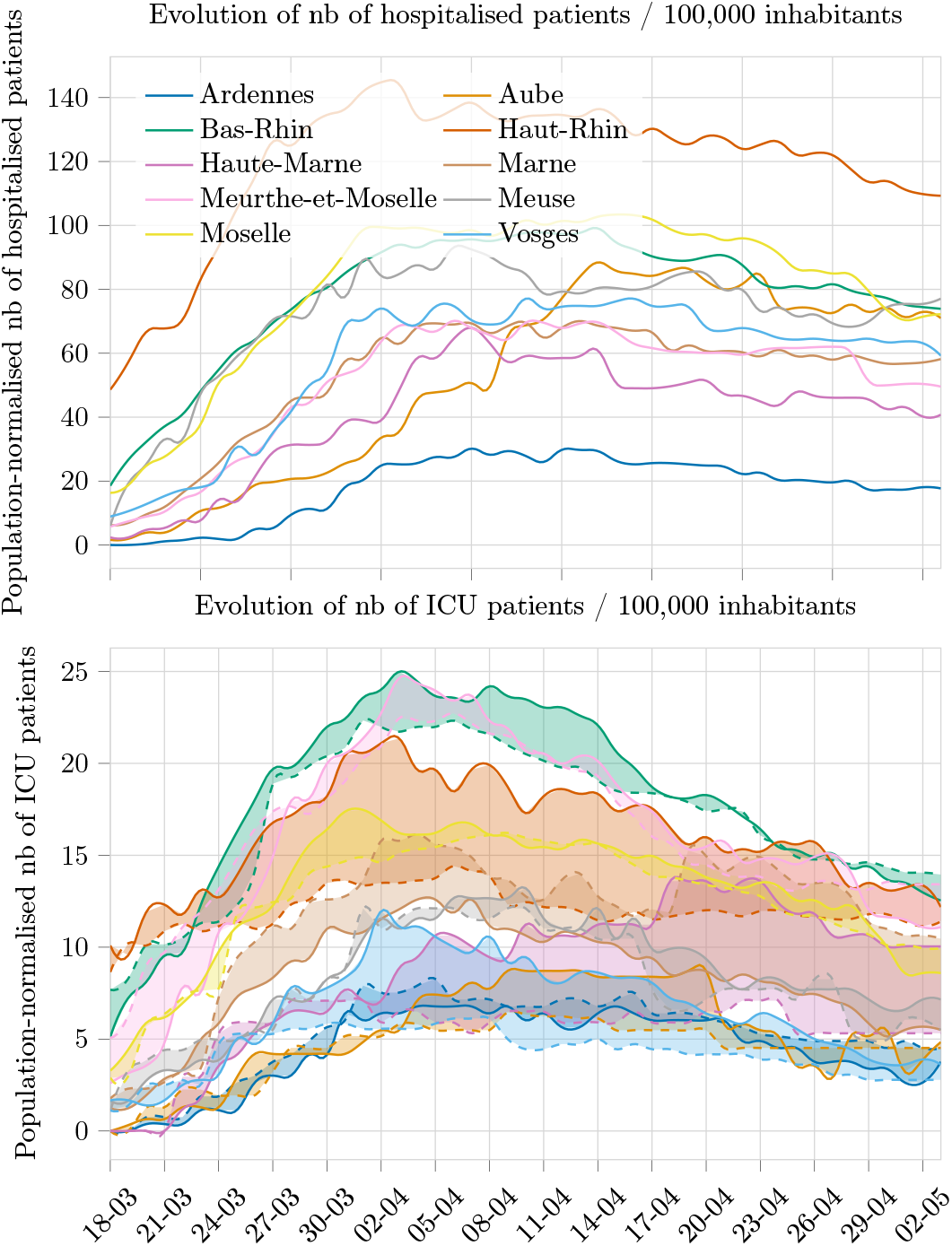
Top: Time evolution of the number of hospitalized patients (official public data Santé Publique France [2020]). Bottom: comparison of the evolution of the number of ICU patients according to the official public data Santé Publique France [2020] (dotted lines) and the `ICUBAM` data (continuous lines).

The discrepancy in the number of patients in resuscitation at the time of reporting between the `ICUBAM` data and the public data is also visible in figure 16. We have observed that the data from `ICUBAM` could be ‘ahead’ from some sources of information (sometimes by two days) indicating that they better represent the reality of the day.

#### F. SEIR model calibrated on the ICUBAM Grand Est data

##### Modeling approach

The `ICUBAM` data give access, for each day t, to the number of patients occupying an ICU bed, noted here *C*(*t*) (’*C*’ for Critical Care), the number of transfers taking place on that day *t*, and the number of refusals that day *t*. We also know the number of deceased patients, and the number of discharged cases cumulated on day *t*. We only consider the total number of beds released, *X*(*t*) (’*X*’ for eXit) (deaths + discarged cases). One writes ordinary differential equations (ODEs) describing the underlying contagion dynamics. In a given *département*, with population size *N*, at each time there is a number *S*(*t*) of susceptible individuals (*S*(*t*_0_) = *N*), *I*(*t*) of infected individuals (denoting Δ*A*(*t*) the variation of a quantity *A* between day *t* − 1 and day *t*)

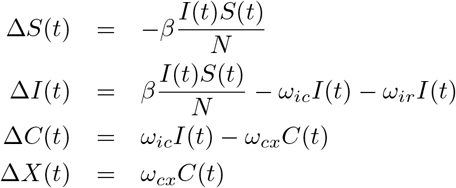

The model used for the figures takes into account an incubation phase (exposed state, SEIR model) and a long exit time, obtained by adding one sub ICU compartment:

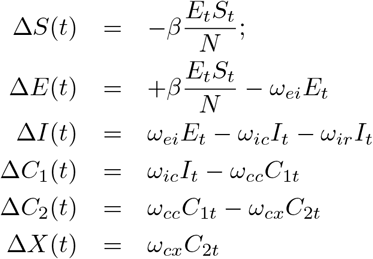

Here the total number of ICU patients is *C_t_* = *C*_1_*_t_* + *C*_2_*_t_*.

Importantly, in the above models we do not consider the refused and transferred patients. One difficulty is that the outcome of these critical care patients initially admitted and thereafter transferred by medical train were unknown. Thus, the models implicitly take into account the local critical care bed capacity. Future work will explore how to incorporate these flows of patients into the modelling approach.

We calibrate all the model parameters in order to have the best agreement between the observed data and the model outcome (with a maximum likelihood criterion, assuming Gaussian noise), the observations being the number of critical care patients and the total number of deceased and discharged alive patients, along the study period.

Since the data do not cover the full epidemic period, we initialize the model by going backward in time: for each *département*, we look for the date *t*_0_ < *t*_1_ such that one gets the best fit by assuming that the epidemic starts at time *t*_0_.

Figures fig. 17 present the results. In Figure fig. 17 (right) we show the Bayesian *credible regions* [Kruschke, 2015] (see below). For the *Aube département*, there was clearly a data entry problem during the first two weeks, with an abrupt entry made at the time of the peak. Remarkably, the observations seem to ‘catch up’ with the model’s prediction. Similarly, for the *Meuse département*, on the last dates the data ‘come back’ onto the model trajectory. Consideration should also be given to the decreasing quality of the data. Thus, for some *départements*, as the *Vosges*, we can observe an inconsistency in the data for the last few days, suggesting a non-entry of departures, while the number of patients in ICU is decreasing. The correct number of patients leaving the ICU is thus slightly increasing in the last days, in better agreement with the model predictions.

**Figure 17:**
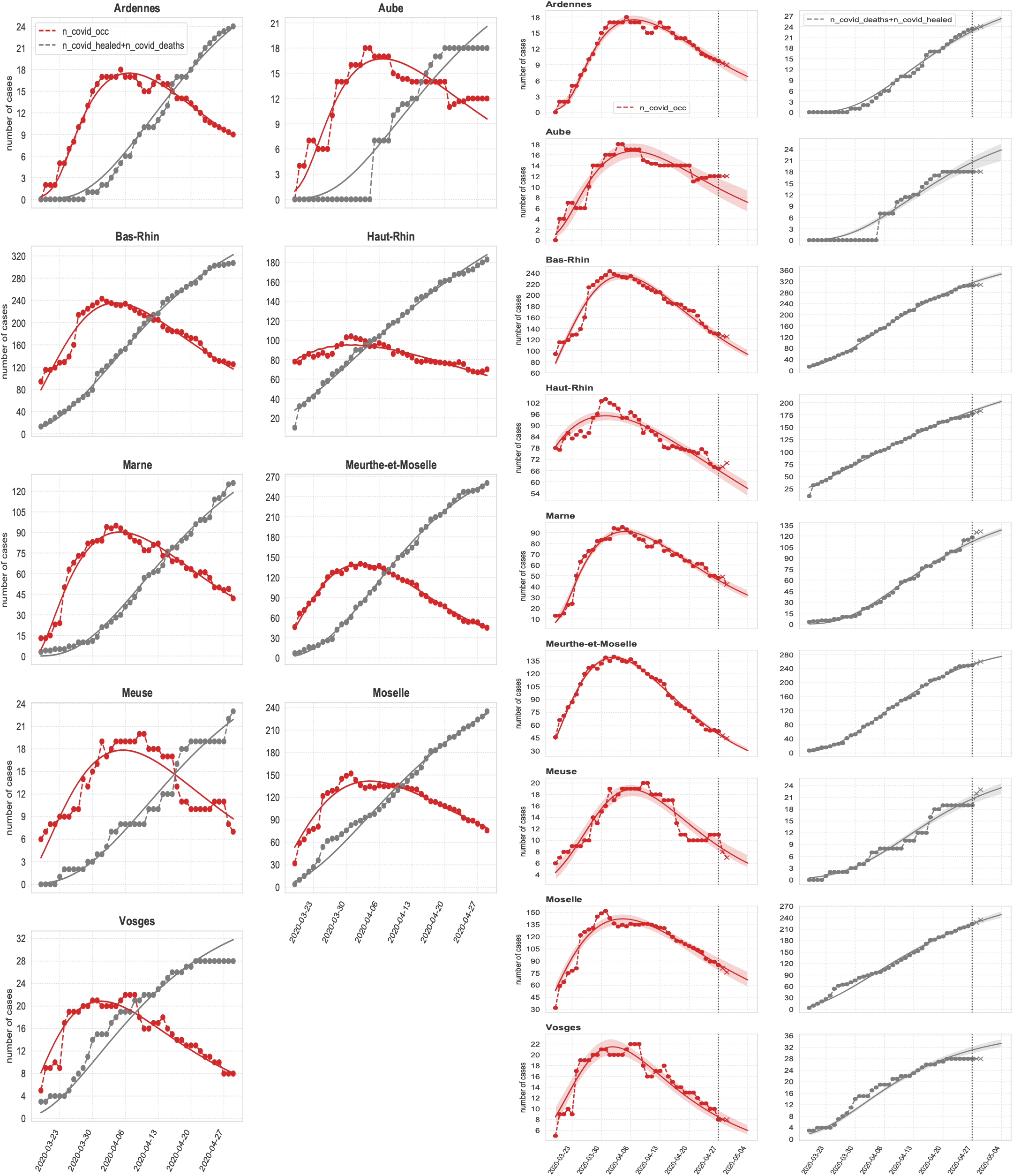
Left: Fits for ICU patients, and the number of (deaths + discharged cases), from the SEIR type model (see text). Model calibrated on `ICUBAM` data as of 29 April 2020. Right: Fits and predictions for ICU patients, model calibrated on `ICUBAM` data as of 27 April 2020 (vertical dotted line; data: circles) and extrapolated for the 4 following days (crosses: data 28 and 29 of April). Red: patients currently in ICU. Black: cumulative number of patients either deceased or discharged from intensive care units. Colored zones (right panel): 95% credible regions (see text).

##### Credible regions

The general idea is the following. Given the observed data, one computes the statistical ensemble of the most plausible scenarii (trajectories) from the Bayesian view point. Let us denote by *χ* the observed data set {*C_t_, X_t_, t* = 1,…,*T*} and Ω = {*β*, *ω_ei_*, *ω_ic_*,…, *ω_cx_*} the set of parameters. We are interested in the posterior *P*(Ω|*χ*) = *P*(*χ*|Ω)*p*(Ω)/*P*(*χ*). The probability *P*(*χ|*Ω) is the result of the model assuming Gaussian noise. *p*(Ω) is the prior on the parameters, and we choose the uniform distribution for every parameter on [0,1]. Given the observed data χ, one can then generate sets of parameters in order to cover 95% of the distribution (more precisely, we consider the 95% highest density region, see [Hyndman, 1996]). This is done with the Slice sampling algorithm (a Markov chain Monte Carlo algorithm method). For each one of these sets of parameters, one generates the associated trajectory 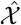. The envelope of these trajectories then defines the 95% credible region. For the variance of the Gaussian noise, we choose a time-independent but *département* specific value estimated from the data.

## Footnotes

1 https://icubam.github.io/about/

2 See Appendix A for a more thorough description of the genesis of `ICUBAM`.

3 Representing a combined population of > 23 million people - See Appendix B for a description of the French administrative divisions.

4 In practice, doctors enter the latest data from their unit in the form in real time, *i.e*. as soon as there is a change.

